# The effects of cannabis use on cognitive function in healthy aging: A systematic scoping review

**DOI:** 10.1101/2020.05.15.20077248

**Authors:** Nina Pocuca, T. Jordan Walter, Arpi Minassian, Jared W. Young, Mark A. Geyer, William Perry

**Affiliations:** University of California San Diego; Center for Stress and Mental Health, Veteran’s Administration San Diego Hospital; Research Service, VA San Diego Healthcare System, San Diego, CA

**Author notes:** Corresponding author. Postal Address: 200 West Arbor Drive, Mailcode 8620, San Diego, CA 92103-8620, USA.

**Keywords:** middle age, older adult, cannabis, ∆9-tetrahydrocannabinol, cannabidiol, cognitive function

## Abstract

Middle-to-older-aged adults (≥50 years) represent the fastest-growing cannabis-using population. Given aging and cannabis use are associated with cognitive decline, it is important to establish the effects of cannabis on cognitive function in this population. This systematic scoping review used PRISMA guidelines to critically examine the extent of literature on this topic and highlight areas for future research. A search of six databases (PubMed, EMBASE, PsycINFO, Web of Science, Family and Society Studies Worldwide, and CINAHL) for articles published by September 2019, yielded 1,014 unique results. Only six articles reported findings for middle-to-older-aged populations (three human and three rodent studies), highlighting the paucity of research. Available human studies revealed largely null results, likely due to several methodological limitations. Nevertheless, the better-controlled rodent studies indicated an age and dose-dependent relationship between ∆9-tetrahydrocannabinol (THC) and cognitive function in aging. Extremely low doses of THC improved cognitive function in very old rodents. Somewhat higher chronic doses were required to improve cognitive function in moderately aged rodents. No studies examined the effects of cannabidiol (CBD) or high-CBD cannabis on cognition. Future research should examine the relevance of age and dose-dependent effects of THC in humans and the effects of CBD on cognitive function in aging.

## 1. Introduction

Middle-to-older-aged adults (≥50 years) comprise the fastest-growing cannabis-using population in Western society (Azofeifa et al., 2016; Fahmy et al., 2012; Han et al., 2017; Kostadinov & Roche, 2017). More than half of middle-to-older-aged US cannabis users report using cannabis medicinally (Choi et al., 2017; Sexton et al., 2019), potentially due to the perception of cannabis as having medicinal effects (Bobitt et al., 2019), a notion perpetuated by dispensaries and popular cannabis websites (Boatwright & Sperry, 2018; Cavazos-Rehg et al., 2019; Luc et al., 2020). This proclivity to use cannabis medicinally likely accounts for why middle-to-older-aged adults tend to select cannabis high in cannabidiol (CBD; the major non-psychoactive cannabis compound), compared to young people, who tend to select cannabis high in ∆9-tetrahydrocannabinol (THC; the main psychoactive cannabis compound; Choi et al., 2017; Sexton et al., 2019). The proliferation of cannabis use among middle-to-older-aged adults may have significant public health consequences, since increases in life expectancy mean that middle-to-older-aged adults are projected to comprise around 40% of the Western population by 2060 (Australian Bureau of Statistics, 2017; Office for National Statistics, 2017; United States Census Bureau, 2018). Of particular concern are the effects of cannabis use on cognition, given this group’s underlying susceptibility to cognitive decline.

Whilst crystalized cognitive functions (e.g., vocabulary) tend to remain stable or strengthen with age (Harada et al., 2013), some middle-to-older-aged adults experience noticeable concurrent declines in fluid cognitive functions including memory, learning, inhibition, attention, decision making, cognitive flexibility, and processing speed (Eppinger et al., 2011; Fraundorf et al., 2019; Kray & Lindenberger, 2000; Marschner et al., 2005; Mell et al., 2005; Samson & Barnes, 2013; Tucker-Drob et al., 2019; Weiler et al., 2008). Significant heterogeneity exists among individuals regarding the degree and extent of age-related cognitive decline (de Frias et al., 2007). Nonetheless, cognitive decline has significant, negative effects for mental health and wellbeing (Burholt et al., 2016; Hill et al., 2016; Parikh et al., 2015; Wilson et al., 2013), underscoring the need to examine factors that may exacerbate age-related cognitive decline. One such factor may be cannabis use, given the known detrimental effects of cannabis on cognition in younger populations.

Cannabis impairs cognition in young people, likely attributable to the detrimental effects of THC on the developing brain (Broyd et al., 2016; Crane et al., 2013; Gorey et al., 2019; Scott et al., 2018). The effects of cannabis use on cognition in middle-to-older-aged adults may be complicated however, by a number of age-related factors, including: (1) An increased selection of high-CBD cannabis, which has anti-inflammatory properties (Burstein, 2015; Mori et al., 2017) and may attenuate the cognitive impacts of low-grade inflammation seen in aging (Fard & Stough, 2019; Patterson, 2015); (2) a slowing of the metabolism, resulting in extended periods of intoxication (Sagar & Gruber, 2018); and (3) age-related changes in the dopamine system (Karrer et al., 2017), which is instrumental in several cognitive domains affected by age including reward-based decision making (Berry et al., 2019), and affected by cannabis use (Yoo et al., 2019). Ultimately, the proliferation of cannabis use among middle-to-older-aged adults who may already be susceptible to cognitive decline and the known detrimental effects of cannabis on cognitive function in young people, highlights the need to examine the effects of cannabis use on cognitive function among middle-to-older-aged adults.

Recent reviews of animal studies found the effect of THC on cognitive function to be age-dependent. Whilst even low doses were detrimental in younger rodents, THC exerted pro-cognitive effects on memory and learning in older populations (Calabrese & Rubio-Casillas 2018; Sarne 2019). Another recent review provided an important overview of some studies examining the effects of cannabis use on aging, with a focus on molecular systems and some consideration of cognition (Yoo et al., 2019). Nevertheless, a systematic review of existing literature, more detailed synthesis and critical appraisal of cannabis effects on aging and cognition across species would ensure that all relevant studies are captured, and provide crucial direction for future research on this emerging issue. Resultantly, the aim of this systematic scoping review is to determine the current extent of the literature, summarize available findings, and identify gaps in knowledge regarding the effects of cannabis use on cognitive function in healthy aging.

## 2. Method

This systematic scoping review was conducted in line with the Preferred Reporting Items for Systematic reviews and Meta-Analyses (PRISMA) guidelines (Moher et al., 2009; Tricco et al., 2018).

### 2.1 Eligibility Criteria

Papers published by September 2019 were included if they examined the effects of whole plant or phytocannabinoids (THC or CBD) on cognitive function in healthy, middle-to-older-aged adult humans (≥50 years) or animals (e.g., mice ≥12 months). No restrictions were placed on publication date or study design. Studies had to be in English and include a baseline or comparison group not exposed to cannabis. Studies that focused exclusively on populations with underlying pathology or substance use disorders (other than cannabis use disorder), or that conflated cannabis with other substances (e.g., polysubstance use), were excluded. Missing or unclear data were clarified by emailing corresponding authors.

### 2.2 Information Sources

Six large databases (PubMed, EMBASE, PsycINFO, Web of Science, Family and Society Studies Worldwide, and CINAHL) were searched using a specialized search strategy, developed with a librarian experienced in systematic reviews (see Supplemental files for the full PubMed electronic search strategy). To ensure saturation of the literature, additional publications were identified via: (1) Grey literature (i.e., research not published as a peer-reviewed article), including conference abstracts and dissertations (searched via ProQuest); and (2) reference lists of included articles and relevant reviews.

Results were exported to Rayyan QCRI, an online service for systematic reviews. Title and abstracts and eligible full text pdfs were independently reviewed, and data for each included study were independently extracted by two reviewers (NP and TJW), blind to each other’s decisions. Disagreements between reviewers were resolved through consensus following each review stage. Extracted data included: participants (sample size, age range, mean age, and sex); exposure (cannabis use or administration definition); comparison group (definition of control/ reference group); outcome information (measures); and main findings.

## 3. Results

The search resulted in 1,014 unique articles for title and abstract screening, yielding 134 articles for full-text review, leading to six articles included in the review (see Figure 1 for the PRISMA flow chart).

**Figure 1.**
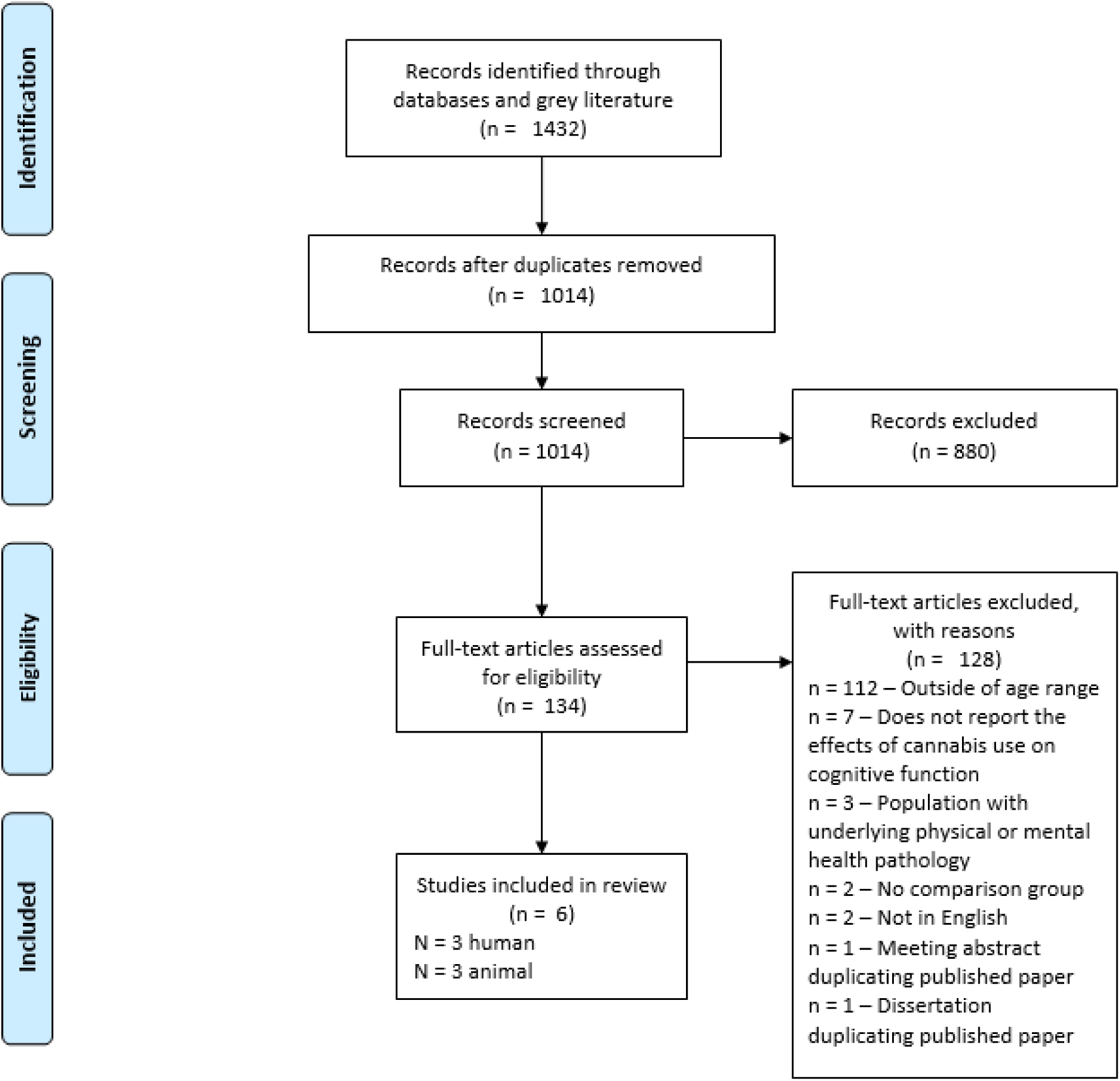
PRISMA Flow Chart

### 3.1 Human Studies

Table 1 outlines the details of the human studies. Most studies (*n*=110;87%) were excluded at full-text review because they did not report results for middle-to-older-aged adults, resulting in three studies and a collective sample of 3,462 cannabis users and 7,917 controls (never used cannabis). The definition of cannabis use differed among studies (i.e., lifetime, past 12-month, or former heavy use). Only Thayer et al. (2019) reported route of administration (i.e., 57% smoked, 32% used edibles, and 11% used both) and cannabis strain preference (although a majority of participants-71%-were unsure of strain characteristics). All studies used computer-based assessments of cognitive functions including working, episodic, and delayed memory, vocabulary knowledge, oral reading skills, cognitive flexibility, processing speed, reaction time, and learning. Burggren et al. (2018) also assessed clinician-reported cognitive function via the Mini-Mental State Examination (MMSE). Only Dregan et al. (2012) found a significant effect of cannabis use on cognitive function. Namely, lifetime cannabis use at age 42 was associated with better memory and executive function at age 50.

**Table 1.** Human studies examining the relationship between cannabis use and cognitive function in middle-to-older-aged adults (≥50 years)

| Author | Sample characteristics | Cannabis use | Cognitive task/s | Main findings |
| --- | --- | --- | --- | --- |
| Thayer et al., 2019 | $N = 56^a$<br>**Cannabis users:** $n = 28$; $Mage = 66.79$, $SD = 5.28$; range = 60–80; females = 10 (36%)<br>**Controls:** $n = 28$; $Mage = 69.79$, $SD = 5.71$, range = 61–83; females = 17 (61%) | ≥weekly for ≥past year | NIH Toolbox Cognition Battery - Flanker inhibitory control and attention test; picture sequence memory test; list sorting working memory test; picture vocabulary test; oral reading recognition test; dimensional change card sorting test; pattern comparison processing speed test | No significant difference in cognitive performance between cannabis users and controls on any of the cognitive outcomes |
| Burggren et al., 2018 | $N = 50$<br>**Former heavy cannabis users:** $n = 24$; $Mage = 65.4$, range = 58.2–72.6; females = 8 (33%)<br>**Controls:** $n = 26$; $Mage = 67.7$, range = 60.6–74.8; females = 12 (46%) | $\geq 20$ days per month, initiated during adolescence ($< 20$ years), continuing for $\geq 1$ years, with $\leq 2$ uses per month after age 35 | Buschke-Fuld selective reminding test – consistent long-term retrieval and delayed recall; Wechsler Memory Scale-II – Logical Memory I and II and Verbal Paired Associates I and II; Rey–Osterrieth Complex Figure – Delayed Recall; Trailmaking Test – Parts A and B; Stroop Test – Word reading speed and interference; Wechsler Adult Intelligence Scale-III – Digit Symbol; Verbal Fluency FAS and Animal Naming tests; Mini-Mental State Examination (MMSE) | No significant difference in cognitive performance between former heavy cannabis users and controls on any of the cognitive outcomes<br>No group differences were found for the MMSE |
| Dregan et al., 2012 | $N = 11,419$; females = 5,734 (50%); all participants were aged 42 at baseline.<br>**Current cannabis users:** $n = 736$<br>**Lifetime cannabis users:** $n = 2,674$<br>**Controls:** $n = 7,863$ | Current cannabis use: Used in the past 12 months; Lifetime cannabis use: Lifetime use, but not in the past 12 months | Immediate and delayed word recall; animal-naming test; letter-cancellation test | No significant effect of current cannabis use at age 42 and cognitive function at age 50. Lifetime cannabis users at age 42 had significantly better scores on memory (immediate and delayed word recall) and executive function (comprised of the animal-naming test and the letter-cancellation test) at age 50, compared to controls. |

<sup>a</sup>Only 28 cannabis users and 10 controls completed cognitive outcomes Mage = Mean age. SD = Standard deviation.

### 3.2 Animal Studies

The three studies administered different doses of THC (see Table 2), intraperitoneally (either by injection or osmotic minipump). Aso et al. (2016) also administered CBD in a 1:1 ratio with THC. Sarne et al. (2018) used female, while the others used male mice. All studies assessed memory, whilst Bilkei-Gorzo et al. (2017) and Sarne et al. (2018) also assessed learning and flexibility. The THC-only studies found a significant positive effect on cognitive function, whilst co-administration of THC and CBD did not significantly affect cognitive function.

**Table 2.**
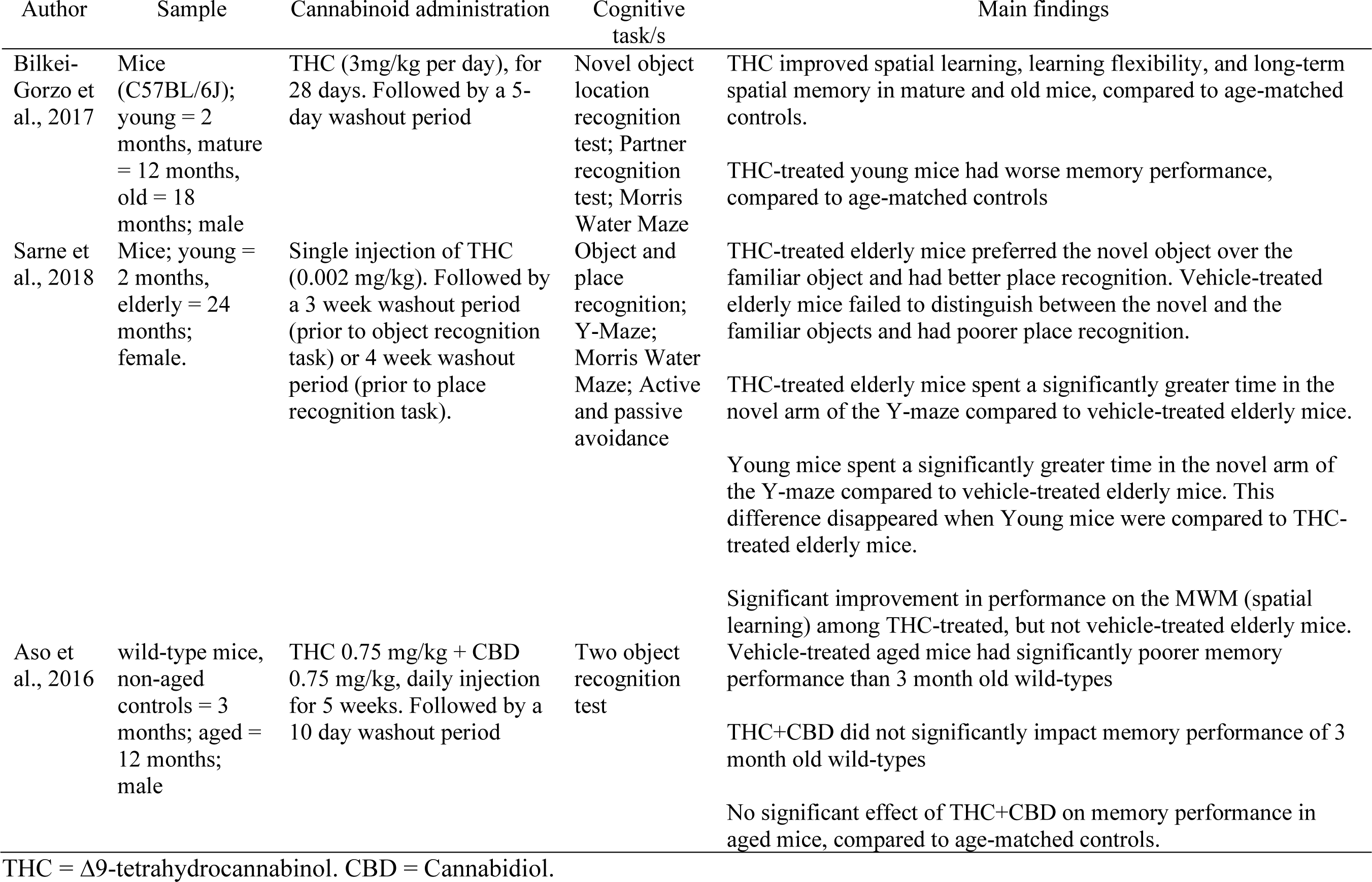
Animal studies examining the relationship between cannabis use and cognitive function in middle-to-older-aged mice (≥12 months)

## 4. Discussion

This systematic scoping review examined current research on the relationship between cannabis and cognitive function in healthy aging. Although a systematic search of six large databases yielded over 1,000 unique results, only six articles satisfied the eligibility criteria for this review, confirming the paucity of research in this area. Most human studies were excluded since they did not report effects for middle-to-older-aged adults. This became apparent at full-text review for most articles, potentially due to the fact that the keyword “middle age” covers 45-64 years. Further, although some studies included a few participants aged ≥50 years (and thus were tagged with the keywords “middle age” or “older adult”), they did not actually report results for this group specifically and therefore were ineligible for this review.

The scant research in this area indicates that existing findings reported herein should be interpreted with caution, since replication and further research are required. Nonetheless, preliminary hypotheses for future research can be gleaned from the reviewed articles. Only one cannabis use variable was associated with cognitive function in humans. Dregan et al. (2012) found lifetime use (≥1 occasions) – but not past 12-month cannabis use – at age 42 predicted better cognitive function eight years later. Although this suggests pro-cognitive effects of cannabis, the relationship may in part be confounded by cognitive reserve, which was not adequately controlled in analyses. Cognitive reserve is a multifaceted construct which buffers against the effects of brain insults, leading to better cognitive function in the presence of brain pathology and normal aging (Satz et al., 2011; Stern, 2009). Future studies should control for cognitive reserve by using a comprehensive measure of the construct and/or a longitudinal approach.

Remaining results indicated that former and current cannabis use were not significantly associated with changes in cognitive function in middle-to-older-aged adults. Burggren et al. (2012) also found no difference between former heavy cannabis users and controls, on memory, attention, processing speed, and executive function in older adulthood. These findings are encouraging and extend the results of the Scott et al. (2018) meta-analysis of 69 studies, which found a remediation of cognitive deficits associated with cannabis use, following 72 hours of abstinence, in young people. Further research is required to examine the effects of early onset (<16 years), chronic cannabis use on cognitive function in later life, given that such use has been linked to poorer cognition in younger populations (Gruber et al., 2012; Sagar et al., 2015). Thayer et al. (2019) found weekly or greater cannabis use was not associated with memory, response inhibition, or processing speed. These null findings may be attributable to the breadth of cannabis use among participants. Examining the effects of cannabis on cognitive function within more homogenous user groups may provide a better understanding of this relationship. This notion of heterogenity impacting interpretable outcomes is supported by the animal studies in this review, which include controlled studies suggesting an age- and dose-dependent relationship between cannabis use and cognitive function.

In contrast to the human studies, a chronic low dose of THC (3 mg/kg/day for 28 days) improved memory, spatial learning, and flexibility in mature and old mice (12 and 18 months; Bilkei-Gorzo et al., 2017), whilst a single, extremely low dose (0.002 mg/kg) improved memory performance and spatial learning in very old mice (24 months; Sarne et al., 2018). Conversely, chronic administration of a 1:1 ratio of THC and CBD (containing 0.75 mg/kg THC, a dose four times less than that administered in Bilkei-Gorzo et al., (2017)), did not affect memory performance in old mice (12 months; Aso et al., 2016). Collectively, these findings underscore age- and dose-dependent relationships between THC, and memory performance and spatial learning, extending the results of Calabrese and Rubio-Casillas (2018). Whilst a single, extremely low dose of THC (0.002 mg/kg) elicits pro-cognitive effects for very old mice (24 months), a larger (but still low) chronic dose (3 mg/kg/day, for 28 days) is required to exert pro-cognitive effects in old mice (12 months). Ultimately, THC may exert pro-cognitive effects by stimulating an endogenous compensatory mechanism, protecting the brain from further insults (Sarne et al., 2018). These animal studies indicate that carefully controlled observational and acute administration studies are required to fully investigate the validity of the hypotheses regarding age- and dose-dependencies of THC effects in human populations. No studies examined the effects of CBD alone, or high-CBD cannabis on cognitive function in aging in animals or humans. Future research should aim to elucidate the contribution of this powerful antioxidant to cognitive function in aging.

The articles examined in this paper have several limitations. Dregan et al. (2012) and Thayer et al. (2019) used self-report to assess abstinence from cannabis, alcohol and other substances prior to cognitive testing; however, they did not confirm self-reported abstinence using biological measures. Thus, participants may have been experiencing intoxication or withdrawal, which have been linked to poorer memory and executive function (Campbell et al., 2017; Crean et al., 2011; Grant et al., 2003; Schreiner & Dunn, 2012). The human studies also did not control for route of administration, which affects THC blood concentrations (Newmeyer et al., 2016). Future research should account for route of administration given the apparent heterogeneity, including high rates of edible use among middle-to-older-aged adults (Sexton et al., 2019; Thayer et al., 2019). The cognitive tasks used in the rodent studies have limited translational validity (Young et al., 2009) and many conflate cognitive function with motor function and novelty preference, which also decrease with age (Bingham et al., 2012; Lindner, 1997; Stansfield & Kirstein, 2006). Further research is also required to examine the effects of cannabis use on dopamine function in middle-to-older-aged adults, given the known effects of cannabis on dopamine function in younger populations (Yoo et al., 2019), and the role of dopamine in general cognition including reward-based decision making (Berry et al., 2019). This potential interaction is also important since age-related changes in the dopaminergic system have been observed (Karrer et al., 2017) and are linked to detrimental mental health and wellbeing outcomes (Volkow et al., 2016; Volkow et al., 2014). Altered dopamine function in middle-to-older-aged adults may explain why older adults tend to be less risk averse than younger populations (Fernandes et al., 2018; Pachur et al., 2017), which may lead to negative practical outcomes (e.g., risky financial behaviors).

## 5. Conclusion

This systematic scoping review underscores a dearth of research examining the effects of cannabis use on cognitive function in middle-to-older-aged adults. Existing human studies have several methodological limitations, potentially accounting for the predominantly null effects. This limitation is exemplified by better-controlled rodent studies that point to age- and dose-dependent relationships between cannabis use and cognitive function in aging. Ultimately, given the recent increase in cannabis use among middle-to-older-aged adults, future human research should examine the relationship between both early and later-life cannabis use on cognitive function within more homogenous, middle-to-older-aged adult cannabis users. This research should also account for confounding factors including acute intoxication or withdrawal, cognitive reserve, route of administration, and cannabis strain preference. Furthermore, further research using animal models are required to determine the mechanistic effects of THC and CBD (together and in insolation) on cognitive function, using measures that account for motor function and novelty preference.

## Acknowledgements

Nina Pocuca is supported by an Interdisciplinary Research Fellowship in NeuroAIDS (IRFN; R25MH081482). Jordan Walter is supported by a T32 fellowship from the NIMH (grant number T32MH018399). This work was supported by the National Institute on Drug Abuse and the Translational Methamphetamine Aids Research Center (TMARC; grant numbers DA043535, P50 DA26306). We would like to thank Karen Heskett for her assistance in developing the search strategy for this review.

